# Community perceptions on effectiveness and safety of indoor residual spraying for malaria control in Uganda

**DOI:** 10.64898/2026.02.05.26345715

**Authors:** Moses Ocan, Nakalembe Loyce, Josephine Bayiga, Geofrey Kinalwa, Emmanuel Arinaitwe, Henry Mawejje, Rogers Naturinda, Moses Adriko, Simon Kigozi, Sam Nsobya

## Abstract

**Background:** Indoor residual spraying (IRS) remains one of the integral tools for malaria control and elimination globally. However, communities have mixed perceptions on the usefulness IRS and other malaria control measures. This study explored community perceptions on the effectiveness and safety of insecticides used in indoor residual spraying for malaria control in Uganda.

**Methods:** A qualitative study employing a phenomenological approach was used to collect data among adult (≥ 18 years) individuals in communities where indoor residual spraying was implemented. Household heads, IRS team leaders, spray operators, community leaders, and IRS field spray supervisors were purposively included in the study. In-depth interviews using an interview guide were used to collect data. Thematic data analysis was done using Nvivo *ver 14*.*0* software.

**Results:** A total of 30 in-depth interviews were conducted. Study participants perceived IRS as effective from the observation of reduction in mosquito population and malaria cases following the spray. Most participants perceived IRS as safe causing only mild effects. The IRS killed other household insects including bed bugs, cockroaches, house flies and ants which helped enhance acceptance. Some of the insecticides used in the spray had strong smell and left stains on the walls. Individuals smeared sprayed walls remove the strong smell and the stains left behind. Participants reported discontinuation of use of bed nets among individuals in sprayed households.

**Conclusions:** The insecticides used in indoor residual spraying were perceived by participants as effective and safe. However, practices common in communities following IRS deployment such as discontinuation of use of bed nets and smearing of sprayed walls could potentially impact the effectiveness of IRS in malaria control. The Ministry of Health and implementing partners should strengthen community education and establish systems to help monitor implementation and effectiveness of indoor residual spraying for malaria control.

## Introduction

Malaria cases in most endemic settings in Africa have remained relatively high albeit slow and unstable reduction in prevalence in some countries(1, 2). The use of multiple interventions including chemoprevention, vector control measures, and current roll-out of malaria vaccines remain the major tools in the fight against malaria(1). Application of IRS has contributed to the elimination of malaria in some malaria affected settings(1). However, in Uganda there is limited evidence on the impact of different malaria control interventions including IRS, mass distribution of bed nets, malaria vaccines and chemoprevention on malaria burden. This is further exacerbated by the dual application of multiple malaria control interventions and the inadequate monitoring and evaluation (3).

In indoor residual spraying, the sprayed insecticide leaves a residue of the insecticide on the interior wall of the sprayed house that is effective to kill mosquitoes thus disrupting malaria parasite transmission (4). A systematic review by Zhou et al., (5) reported lower rates of malaria after IRS application and was associated with over 80% coverage. The World Health Organization (WHO) recommends use of IRS (4) as a proven and effective malaria control measure. However, with the limited monitoring of IRS implementation in most malaria endemic settings especially in sub-Sahara Africa, the impact of IRS and other malaria control interventions remains unknown. This is further exacerbated by the impact of climate change on malaria transmission.

A systematic review by Zhou et al., (5) report that DDT, pyrethroids, methyl carbamate, and combined use of multiple insecticides have high effectiveness in malaria control. Insecticide resistance is emerging to nearly all the malaria mosquito vectors including *Anopheles culicifacies, An. gambiae, An. coluzzii*, and *An. stephensi* across malaria endemic countries (1, 6). Continued susceptibility of the local malaria mosquito vectors is a key determinant of IRS effectiveness. The reported vector resistance against current insecticide threatens to reverse the gains of IRS and insecticide treated bed net distribution in malaria control(1, 6). Factors including timing of IRS deployment and level of training of the spray operators that potentially affects quality of spraying further impacts the effiectiveness of IRS in malaria control (7). This is exacerbated by complex IRS implementation logistics, coupled with community hesitancy in most malaria affected settings especially in Sub-Sharan Africa(1, 8).

Indoor residual spraying (IRS) is the second most widely implemented vector control intervention by most National Malaria control Programs (1). When implemented correctly, IRS has been shown to be a powerful intervention to reduce malaria mosquito vector density and, therefore, to reduce malaria transmission(9). The success of malaria control interventions requires high coverage and utilization at individual and community levels (10). However, coverage of IRS is dependent on the perceived benefits of the intervention and its effectiveness against the malaria vector and extent of undesired side effects (11) which also influence acceptability. Community acceptability is a prerequisite for effective implementation of vector control programmes. In addition, studies have shown that side effects associated with insecticides used for IRS contribute to acceptance or refusal of the intervention (12, 13). This study explored community perceptions on the effectiveness and safety of insecticides used in indoor residual spraying in West Nile region, Uganda. The findings of the study provide context specific evidence which is key for policy and decision makers to help strengthen implementation of IRS for malaria control.

## Materials and methods

### Study design and settings

The study employed a phenomenological approach and was conducted in settings were indoor residual spraying using Clothianidin-deltamethrin (Fludora Fusion) and Pirimiphosmethyl (Actellic) was implemented in West Nile region, Uganda. Data collection was done from February to June 2024.

### Study population, sample size and sampling

Data was collected among consenting adult (≥ 18 years old) participants in communities. The participants included household heads, spray operators, spray team leaders, IRS supervisors and community leaders. Information saturation was used to determine the sample size. Individuals in households that received the spray and those who had participated in the deployment of IRS in the communities were purposively selected for inclusion into the study. Both male and female participants were approached face-to-face for enrolment and for the interviews.

### Data collection procedure

Data was collected using in-depth interviews (IDI) conducted with study participants following an interview guide. The interview guide was developed using information from previous studies (3, 8). The interview guide was pilot tested among community individuals in Tororo district and was also reviewed by staff from integrated vector management of the National malaria control program, Ministry of Health. The findings from the pilot and the feedback from the review were used to adjust the interview guide prior to data collection. The interview questions, prompts and guides covered socio-demographic characteristics, IRS insecticides used, acceptability, effectiveness, adverse effects of IRS insecticides, uptake of IRS, and community practices following spraying.

The in-depth interviews were conducted by two male (OW, TT) and one female (KK) research assistants with Bachelor of social science degree. The lead researcher (OM), a male clinical pharmacologist (PhD) and a faculty member at Makerere University also conducted participant interviews. The three research assistants were community development officers and conversant with the local language. Two research assistants (TT, KK) were residents in communities that received IRS in the study setting. While OW and OM were not from IRS settings in West Nile. The interviews were conducted with participants at a convenient location in homes or work places with no non-participants during the interviews. The interviews were done using a language that the participant was most comfortable with. Probe questions were used to get further details on topics relevant to the research question. In addition, attempts were made by the interviewers to gain narratives from participants about each topic to gain insights into the practicalities and social realities of how the intervention was enacted and perceived by those implementing, supporting, and observing it. The interviews were recorded using an audio recorder (SONY, ICD-PX470, China). In addition, field notes and non-verbal expressions of participants were captured during the interviews. Research assistants conducted repeat interviews among two purposely selected participants for clarification. The interviews took on average 45-60 minutes.

### Data management and analysis

After each interview, short summaries of the discussion including field notes were typed into Microsoft word and shared with the study participants for verification. Audio files were downloaded onto a password protected computer hard drive and backed up regularly. Audio recordings were listened to carefully and then transcribed into Microsoft word in English ready for exporting to NVivo *ver 14*.*0* (QSR International, Cambridge, MA) qualitative data management software for coding and analysis.

Interviews were analysed using a coding scheme developed from pre-defined topics together with themes emerging from the data. Two researchers (BJ, SN) coded the data. Prior to the coding process, a coding tree was developed listing all the emerging themes and sub themes. Each interview was reviewed and all statements pertinent to the sub themes identified, extracted and placed under the identified themes by placing the verbatim statement under a relevant label or theme. The process of labelling all the statements from the data collected (coding) was completed using qualitative data analysis software, NVivo (QSR International, Cambridge, MA). The initial coding was conducted independently by the lead social scientist experienced in qualitative data analysis (BJ), discussed with the principal investigator (MO) leading to a final coding scheme which was agreed upon and applied to all transcripts. Alongside this coding, a reflective analytical diary was kept, to draw out and justify emerging themes and lines of enquiry through the fieldwork process. Findings from interviews were presented as paragraphs of descriptive narratives supported by participant quotes where applicable.

### Ethical considerations

The study proposal was reviewed and approved by the Makerere University School of Biomedical Science Research Ethics Committee (SBS-2023-456). Research clearance to conduct the study in Uganda was obtained from Uganda National Council of Science and Technology (HS341ES). Administrative clearance was also be obtained from study districts and hospitals. A written informed consent was obtained from study participants prior to the interviews.

## Results

### Characteristics of study participants

The in-depth interviews were conducted among 30 respondents including spray operators (9), household heads (7), spray team leaders (7), spray supervisors (5), and community leaders (2). The following themes emerged from data analysis.

1. Suspected adverse effects of insecticides used in indoor residual spraying
2. Community perceptions on effectiveness of insecticides used in indoor residual spraying
3. Persistence of mosquitoes in houses following deployment of indoor residual spraying in communities
4. Practices of individuals in communities following deployment of indoor residual spraying for malaria control

### Suspected adverse effects of insecticides used in indoor residual spraying

The study participants (15/30) reported not to have experienced any serious suspected adverse effect following spray with either sachet (Fludora Fusion) or bottle (Actellic) insecticide. The respondents attributed the absence of adverse effects to adherence to the instructions given by the spray teams.

> *“In my family, this sachet chemical spray (Fludora Fusion) actually …. had no bad effects unless you come into contact immediately. On ourselves, it didn’t have much effect*… *because we tried to follow what we were told by the sprayers”* --IDI-07-Household Head.
>
> *“…there is no reaction on people except only that on a sprayer on the face that is what we have seen…*.*” -*--IDI-19-Spray Operator.

Some participants reported minor adverse effects of the insecticides that were used in IRS including itching of the skin and eye, difficulty in breathing, headache, and skin rash.

> *“……during the spraying, you could find you experience headache, [Mm] the itchiness of the body, sometimes around the neck, plus the face” --*IDI-21-Team Leader of Spray Operators.
>
> “*But again, the challenge I realized was the sachet chemical (Fludora) actually …. generates heat. It generates heat and if you happen to come into contact with it when it is still wet on the walls it causes itching on the body*…*”* ---IDI-07-Household Head

The IDI participants reported that the bottle chemical (Actellic) had a bad smell unlike the sachet insecticide (Fludora Fusion) and was generally bad for everyone however, pregnant women were more affected.

> *“The complaint was mostly with the pregnant women. For them they were complaining that the spray makes their heads to ache, and cause vomiting …” -*--IDI-02-Household Head.

### Community perception on effectiveness of insecticides used in indoor residual spraying

The respondents reported on perceived effectiveness of the insecticides used for IRS in malaria control. Study participants mentioned that the mosquitoes disappeared from the house after some time following the spray. However, other respondents shared that the mosquitoes were seen coming from outside the sprayed rooms. Some participants said household members experienced the comfort of sleeping without a net because the mosquitoes disappeared following spraying.

> “*So, the first day I couldn’t sleep properly because the room was all wet with the spray…and also I was hearing sounds of the mosquitoes* ***[mm]*** *then after some times, the sound disappeared completely*… *this was something which was very unique for me*” ---IDI-02-Household Head
>
> *“After spraying, people didn’t experience the mosquitoes in their houses. And it has taken some time for mosquitoes to again come to those houses. And it has also reduced the burden of always taking children to hospital because of malaria*…*” --* IDI-04-Community Leader
>
> *“All in all, the two chemicals (Sachet and bottle) are okay…they are working*…*” ---* IDI-24-Supervisor.

Study participants reported that malaria cases which were rampant among infants in households had reduced following the spray.

> *“But after spraying, I’ve seen some change the cases of malaria have reduced. So, it used to be like in a day a community health worker would see children sick of malaria like three, or four in but these days after the spray there is change…”* --IDI-03-Team Leader of Spray Operators.
>
> “*After spraying, malaria cases have reduced in my home it’s not like the period before spraying. I see there is a difference*” ---IDI-25-Household Head.

Malaria is one of the leading causes of morbidity and mortality in children under five years and causes significant economic strain to households especially on out-of-pocket expenditure on healthcare. Thus, the observed reduction in malaria cases in homes brought financial relief to families with one respondent reporting that the money they used to spend on treating malaria can now be used for other activities.

> *“… the money that I used to spend on malaria treatment is no longer happening. I can now divert the little I have to do other things*” --IDI-02-Household Head.

Collateral effect of IRS on other household insects further enhanced the perception of its effectiveness. Study participants reported that IRS killed most insects in the house including house flies, cockroaches, bed bugs, and ants. Despite the promotion of IRS for malaria control by government and the implementing partners, the colleterial effect on other house insects potentially help motivate communities to accept IRS.

> “…*the cockroaches, houseflies, bed bugs and all other insects in the house died immediately from the spray…the 2022 chemical (Fludora Fussion) killed a lot of the insects in the house…”*
>
> *---IDI-12-Supervisor*

### Persistence of mosquitoes in houses following deployment of indoor residual spraying in communities

Some respondents reported that for the sachet IRS (Fludora Fusion), it took some time after the spray for the mosquitoes to disappear from the house. The participants reported that the mosquitoes could still be seen and kept making noise in the house after spraying.

> “*The mosquito bites continued after the spray with the one in the sachet (Fludora Fusion)… so meaning though my house was sprayed but I believe that it did not kill mosquitoes…*.*”* ---IDI-25-Household Head

There were several expressions of doubt about the quality of mixing of the spray using the insecticide in the sachet (Fludora Fusion) and how it worked. When participants saw mosquitoes in their houses, they considered the insecticide as not effective. The participants could not gauge if the operators were mixing the right quantities of the insecticide in the spray tanks. The extra flying and/or noise from the mosquitoes reported in the sprayed houses is potentially due to the excito-repellency effect of Deltamethrin a component of Fludora Fusion spray insecticide.

> *“…. Iam getting into this house and these mosquitoes are also there. Why? Did you really mix the chemical in a good way…how come that mosquitoes are not dead, Why?”* --IDI-20-Spray Operator.
>
> “*That time in 2022 there was a variation because others were saying, these people [sprayers]did not mix as well*… *now this was all about lack of Education…the whole thing was confusing us. But in 2023 (bottle IRS) there was now experience with IRS in the communities*” ---IDI-02-Household Head.

Some participants reported that the mosquitoes were seasonal and that it may be hard to determine if the spraying worked or the season of mosquitoes is at the peak. While mosquito population and cases of malaria reduced following IRS, the effect was short lived, and this caused frustration among individuals in communities.

> “*The reduction is just for some times, but after here we begin realizing the cases again… after a year when it starts to rain, we shall begin realizing mosquitoes and many cases of malaria*” ---IDI-02-Household Head.

The reason given by study participants for the persistence of mosquitoes in houses was the practice of smearing sprayed walls of houses which is a common practice in rural communities.

> *… for others that smell appeared to be something which was very bad to them. So, they took off the smell by smearing the wall, and those are the people who are complaining that they are having mosquitoes in their rooms*… *But the ones who smeared their walls off the chemical had mosquitoes in their rooms*,” --IDI-02-Household Head

### Practices of individuals in communities following deployment of indoor residual spraying for malaria control

The study participants reported different practices among individuals in sprayed houses which could potentially impact effectiveness of IRS for malaria control. These practices are key in understanding community dynamics for future IRS spray programs. Individuals in communities continued sleeping in their sprayed houses.

> “…*that same day the house was sprayed is when I cameback and we slept inside the house since it was already sprayed…” --*IDI-07-Household Head.
>
> *“Yes, they slept in the sprayed house … we told them that after spraying the house is safe…*” --IDI-09-Spray Operator.

The IDI participants reported that some people smeared their houses before the spray exercise after learning that they would not be allowed to smear the walls of their houses for some time after the deployment of IRS. Some people smeared their houses before the spray due to sensitization against smearing the walls of the house after spraying.

> “…*We told them, you people you have to smear all the houses before the spray… they got the information before the spray*” --IDI-08-Team Leader of Spray Operators.
>
> *“We got that, … information before [Mmm] that we have to smear our house before spraying [Mmm] and all of our houses were smeared before…” -*--IDI-14-Household Head.

In communities after deployment of IRS, the study participants reported that individuals in households felt that after the spray, mosquitoes had disappeared, so they stopped sleeping under the bed nets.

> *“After spraying the houses… many people refused to use their bed nets, thinking that spraying will protect them from mosquitoes*… *even up to now, if you go to other homes, they are not using their nets. They have packed their bed nets with that mindset that after spraying no more malaria”-*--IDI-02-Household Head.

The study participants reported complaints of mosquitoes biting individuals outside the sprayed house. With cases of delays in entering the sprayed houses in the evening among individuals in the communities, the risk of getting malaria from mosquito bites outside the sprayed house is more likely. Indoor residual spraying targets indoor resting malaria mosquitoes however, the change in vector behavior and resistance in IRS settings could impact the effectiveness of IRS in malaria control.

> “*The mosquitoes do not only bite us from inside the house. When you’re outside, they can still bite you and you can still get the malaria…”* ---IDI-07-Household Head.

Participants shared that people spend a lot of time in the evening in outdoor social events including watching cinema, playing cards, and taking drugs like khat.

> “*They can stay outside for long the mosquito can also bite them. The cards, playing cards, Chewing khat (canabis). At that moment, the mosquito can even bite your body*” ---DI-09-Spray Operator.

## Discussion

Indoor residual spraying (IRS) remains the cornerstone in malaria elimination efforts however, recent global trends in health financing threaten scaling up of its implementation especially in high burden sub-Sahara Africa countries (1). Additionally, insecticide resistance and change in vector behavior further threaten the role of IRS in malaria control (1, 6). The IDI participants in this study reported increase in outside biting and delay in clearing of mosquitos in sprayed households. This is consistent with the findings of a previous study in northern Uganda by Mwesige et al., (6) that reported change in vector composition and behavior in IRS settings. These observations by communities were perceived as ineffectiveness of IRS in malaria control. There were mixed perceptions on IRS effectiveness in communities, most of the study participants reported decrease in mosquito population and reduction in malaria cases following IRS. The insecticides also had collateral effect on other house insects a further indicator of IRS effectiveness. These mixed perceptions by individuals on IRS effectiveness could be an indicator of inadequate community education. While perception on IRS effectiveness remains a key driver of acceptance and uptake (13), there is need for the malaria control programs to prioritize community education to prevent misinformation on IRS. However, the perceived effectiveness of indoor residual spraying using the two insecticides (Clothianidin-Deltamethrin, Fludora Fusion and Pirimiphos-methyl, Actellic) provides opportunity for scaling up IRS in malaria elimination efforts.

In this study participants reported no major adverse effects from the insecticides used in IRS however, the perception that the insecticides were dangerous to human health persisted. Some of the reported effects included itching of the skin and eye, respiratory distress, headache, and skin rash. This is consistent with the findings of previous studies (12, 14) that reported concerns on the side effects of IRS in communities. Experience of unwanted effects among individuals in communities is likely to influence acceptance of IRS. A previous study by Magaco et al., (13) in Mozambique found that negative experiences with IRS was one of the primary barriers to uptake. The reported effects also occurred among the spray operators and could be due to limited personal protective equipment (PPE). The inadequate supplies for the spray operators reported in this study remains a limitation for IRS deployment in most malaria endemic settings in Africa.

In most houses following IRS deployment and subsequent reduction in mosquito population, majority of the individuals stopped using bed nets. Additionally, study participants reported practices such as smearing of sprayed walls and delay in entering houses in the evening. This is unlike findings of previous studies (13, 15, 16) in Mozambique which reported that individuals in communities preferred use of ITNs over IRS for malaria control. Perception of the effectiveness of IRS against mosquitos could have enhanced the practice of stopping the use of bed nets reported in this study. However, in Mozambique communities preferred ITNs as they were perceived to be more effective compared to IRS (13). Although there is limited data on the effectiveness of deployment of multiple malaria control measures, adherence to all the interventions remains key for malaria control. This is especially the case due to the complex and changing behavior of both mosquito vectors like outdoor biting and humans including late night outdoor activities (1, 6). The variation in community perception of the effectiveness of the main malaria vector control measures, IRS and ITN is an indicator of the need to strengthen monitoring, community education and social behavior change communication in the implementation of malaria control interventions.

The smell of the insecticides used in IRS was perceived by communities as an indicator of its effectiveness in malaria control. Some of the study participants indicated that it was the smell of the insecticides that chased and killed the mosquitoes. Additionally, the smell was used as a sign to indicate that the house was sprayed. However, most study participants attributed the strong smell of the insecticide especially Actellic to side effects of IRS. In response to the strong smell and the stain left behind by the insecticides used in IRS, most individuals smeared the sprayed walls with clay or cow dung while others covered the walls with clothes/curtains. This could potentially impact the effectiveness of IRS on malaria control in communities. As reported in our study where study respondents indicated that individuals who smeared their walls were the ones complaining of persistence of mosquitoes in their houses following the spray. This is like findings of a previous study in Mozambique which found the strong smell and stain left behind by the insecticides as a barrier to IRS implementation (13).

The study had some limitations for instance the first round of indoor residual spraying was in 2022 therefore recall bias is possible. However, being a major event that was done for the first time and addition to the use of multiple key stakeholders involved in the IRS exercise the responses were corroborated between different participants and thus ensuring consistency.

## Conclusions

The insecticides used in IRS in West Nile region were perceived by participants as effective and safe. However, practices common in communities following IRS deployment such as discontinuation of use of bed nets and smearing of sprayed walls could potentially threaten the effectiveness of IRS in malaria control. The Ministry of health and the implementing partners should strengthen community education and establish systems to monitor implementation and tracking of IRS effectiveness.

## Data Availability

All study data are available within the manuscript

## Acknowledgments

The efforts of Olwortho Wilfred and Obeida Tabuga the social scientists who conducted field interviews for collection of data for the study. We also acknowledge the study participants and district leaders who help in enabling the study team access individual participants in communities for the in-depth interviews.

